# Statistical refinement of case vignettes for digital health research

**DOI:** 10.1101/2024.08.30.24312824

**Authors:** Marvin Kopka, Markus A. Feufel

## Abstract

Digital health research often relies on case vignettes (descriptions of fictitious or real patients) to navigate ethical and practical challenges. Despite their utility, the quality and lack of standardization of these vignettes has often been criticized, especially in studies on symptom-assessment applications (SAAs) and triage decision-making. To address this, our paper introduces a method to refine an existing set of vignettes, drawing on principles from classical test theory. First, we removed any vignette with an item difficulty of zero and an item-total correlation below zero. Second, we stratified the remaining vignettes to reflect the natural base rates of symptoms that SAAs are typically approached with, selecting those vignettes with the highest item-total correlation in each quota. Although this two-step procedure reduced the size of the original vignette set by 40%, comparing triage performance on the reduced and the original vignette sets, we found a strong correlation (r = 0.747 to r = 0.997, p < .001). This indicates that using our refinement method helps identifying vignettes with high predictive power of an agent’s triage performance while simultaneously increasing cost-efficiency of vignette-based evaluation studies. This might ultimately lead to higher research quality and more reliable results.

## Introduction

In the field of digital health research, case vignettes – that involve either fictitious or real medical scenarios – have become a widely accepted methodology (1–6). The reliance on vignettes is primarily due to practical constraints: direct involvement of patients may often be infeasible or unethical, comparability across patients can be limited, and specific research scenarios may present additional barriers to using real patients (7,8). To mitigate these constraints, researchers frequently use case vignettes as proxies to conduct these studies. However, the vignettes used in digital health research are often developed in an unstandardized way and without theoretical foundation (9–12).

Particularly in research focused on symptom-assessment applications (SAAs), many studies have adopted a set of vignettes developed by Semigran et al. in 2015 (9). This vignette set – which was derived from diverse medical resources – has not only been used in studies examining the triage performance of SAAs and laypeople but also in the evaluation of large language models (LLMs) (13–16). Although these vignettes marked a significant step in evaluating SAAs, they have also been criticized. Key concerns include the development process (e.g., scenarios based on medical textbooks may not reflect ecologically valid descriptions of real patients), and the lacking validation of these vignettes (i.e., all developed variables are used without any quality assessment) (11,12,17). This criticism raised not only questions about the suitability of vignettes for accurately estimating the triage performance of human and digital agents, but also whether some vignettes might be better suited for evaluations than others (12). For example, it may be unclear which cases are easier or more difficult to solve and whether vignettes have incremental predictive power or could be omitted. It is also unclear if the predictive power of a vignette differs between human and digital agents and if different vignette sets might be needed for each agent. Thus, it is no surprise that Painter et al. recommend creating guidelines to identify which vignettes to include in accuracy evaluations (11).

The problems associated with existing vignettes highlight an urgent need for a more systematic and theory-driven approach to developing case vignettes. Ecological psychology and test theory provide a framework for addressing these challenges (18). By applying test-theoretical approaches, researchers can identify vignettes with high predictive power for assessing the performance of SAAs and other digital tools. In a previous study, we have shown that test-theoretic metrics can be readily applied to case vignettes in digital health research and that the current sets of vignettes (e.g., the one suggested by Semigran et al. (9)) are problematic in this regard (12). In another study we have outlined a method based on Egon Brunswik’s concept of representative design to develop vignettes with high ecological validity (19).

The current paper takes these efforts a step further by detailing how to refine any existing vignette set using test-theoretical metrics. This refined set aims to make case vignettes studies more cost-efficient by reducing the number of vignettes required while simultaneously identifying and maintaining the vignettes that most effectively predict the performance of different diagnostic agents.

## Method

### Study Design

This study presents a two-step procedure based on test theory to refine an existing set of case vignettes to test the triage performance of different agents. Specifically, our goal is to refine the full set and arrive at a validated subset of vignettes for each agent. To validate the presented vignette-refinement procedure, we compare the performance of the original and the refined set of vignettes based on data collected from laypeople, SAAs and LLMs in a previous study (19).

### The Original Vignette Set and Data

The original vignettes were developed with the RepVig Framework according to the principles of representative design as outlined in Kopka et al. (19). That is, the vignettes were selected through random sampling of actual patient descriptions, where individuals presented symptoms in their own words and asked whether and where to seek medical care. The full vignette set was developed to reflect the natural base rate of symptoms that SAAs are approached with, using symptom clusters for stratification as reported by Arellano Carmona et al. (20).

A total of 198 laypeople (with no medical training) evaluated the urgency of these case vignettes (20 vignettes each, resulting in a total of 3,960 assessments). Additionally, the dataset encompasses evaluations from 13 SAAs, each tested across all vignettes by two research assistants without a professional medical background. Furthermore, for LLMs, the lead author collected data on five LLMs that were openly available and offered a chat interface.

### Statistical Refinement

To refine the vignettes, we applied test-theoretical metrics in a first step: *item difficulty* (ID, how difficulty vignettes were to solve for any one agent) and *item-total-correlations* (ITC, how solving a given vignette correlates with solving other vignettes of the same acuity) (12,21). For each agent, vignettes with an ID of zero were excluded because they were unsolvable for the corresponding agent, and thus offered no insight into differences in triage performance. Similarly, vignettes with negative ITC values were removed because they negatively correlate with triage performance, rendering them unsuitable for performance evaluation according to test theory standards (21).

In a refined vignette set, the natural base rates reflected in the original set should be maintained to ensure that the distribution of symptom clusters still reflects real-world conditions, preventing any one cluster from disproportionately influencing evaluation results. In a second step, we thus evaluated the relative proportion of the smallest remaining symptom cluster and adjusted all other clusters to match this proportion to ensure that the base rates in the refined vignette set remained unaltered. For instance, if the smallest quota retained 3 out of 5 vignettes, this corresponds to 60% of the original size. Consequently, we adjusted the size of all other quotas to reflect this proportion and included 60% of vignettes in the other quotas as well. In each quota, we retained the vignettes with the highest ITCs as these have the highest predictive power.

### Validation

Finally, to validate the refined set, we compared the degree of association between performance estimates derived from the subset with those derived from the full set. We used common metrics for triage performance evaluations: accuracy, accuracy for each triage level, safety of advice, inclination to overtriage, and – this metric can only be calculated for SAAs – the capability comparison score, which is a score that allows capability comparisons between SAAs that were tested with differing vignette sets (12). We calculated these metrics using the symptomcheckR package (22) for every person, every SAA, and every LLM using both the full and the refined vignette sets and assessed the degree of similarity between these outcomes using Pearson correlation. Following a review by Overholser and Sowinski (23), we interpreted a correlation above 0.90 as very high, between 0.70 and 0.90 as high, between 0.40 and 0.70 as moderate and below 0.40 as low or negligible.

## Results

### Refinement

Step 1 for laypeople: All (45/45) vignettes from the full vignette set have an ID greater than zero. Thus, no vignettes were excluded because of the ID. Six vignettes have an ITC below zero and were excluded, see Figure 1.

**Figure 1.**
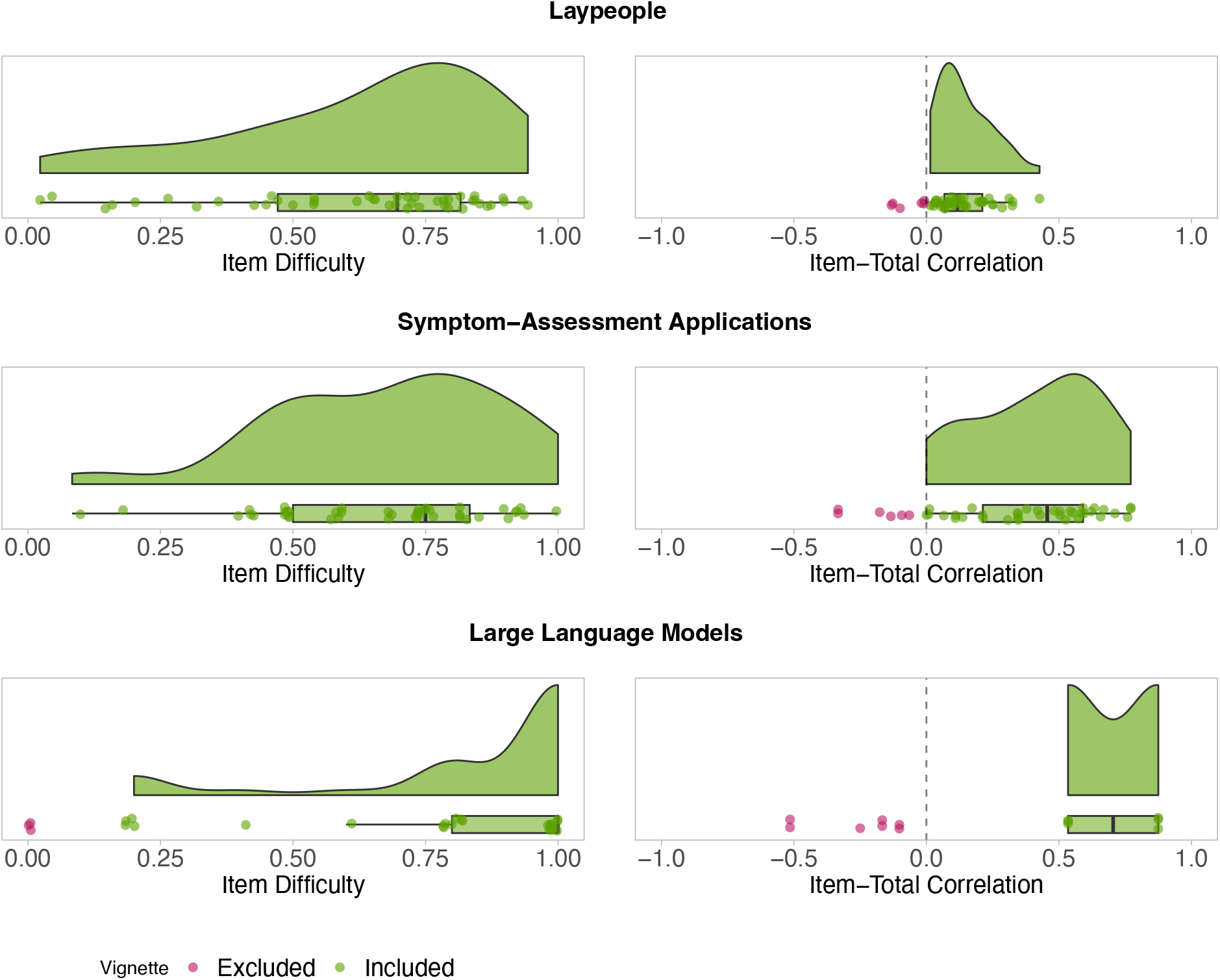
ID (left) and ITC (right) for each full vignette and agent. Red points represent vignettes that do not satisfy test-theoretic criteria and were excluded. Green points show vignettes that are included in the refined vignette set.

Step 2 for laypeople: The biggest reduction occurred within the “other pain” symptom cluster, which was reduced from 5 cases to 3 cases. This reduction corresponds to a 60% retention rate. Consequently, we adjusted the size of all quotas to reflect this new size, reducing them to 60% of their original sizes. The new quota sizes are shown in Table 1. In each symptom cluster, these quotas are filled beginning with the vignettes that have the highest ITC. For example, in the cluster “musculoskeletal pain”, this corresponds to the vignettes 1 (ITC_1_ = 0.312), 4 (ITC_4_ = 0.249), and 2 (ITC_2_ = 0.204), as the other vignettes had lower ITCs with ITC_5_ = 0.151 and ITC_3_ = 0.064. Clusters that only had 1 vignette originally (n = 3 clusters) are joined together and the two vignettes with the highest ITC in this joined cluster remain in the filtered set. The refined vignette set can be found in the Appendix.

**Table 1.** New quotas for refined vignette sets for laypeople and SAAs after excluding vignettes that did not satisfy test-theoretic criteria. The clusters are based on Arellano Carmona et al. (20).

| Symptom Cluster | # of vignettes in full set | # of vignettes in refined set |
| --- | --- | --- |
| Musculoskeletal Pain | 5 | 3 |
| Joint Pain | 3 | 2 |
| Headache | 1 | <1, joined with other clusters |
| Chest Pain | 1 | <1, joined with other clusters |
| Other Pain | 5 | 3 |
| Gynecological | 5 | 3 |
| Tumors/lumps/masses | 4 | 2 |
| Edema | 2 | 1 |
| Skin issues | 2 | 1 |
| Gastrointestinal | 2 | 1 |
| Impaired sensations | 3 | 2 |
| Urinary Tract Problems | 3 | 2 |
| Upper Respiratory Symptoms | 1 | <1, joined with other clusters |
| Other | 8 | 5 |

Step 1 for SAAs: All (45/45) vignettes have an ID greater than zero and remained in the set, while six vignettes had an ITC below zero, see Figure 1. These cases were excluded:

Step 2 for SAAs: :The biggest reduction (2 vignettes) occurred in the “other pain” symptom cluster again. Because this represents the smallest new cluster now, all quotas are reduced to 60% of their original size (with the same retention rate as for laypeople, see Table 1). Those quotas are filled again with vignettes with the highest ITC in each cluster. For “musculoskeletal pain”, this corresponds to the vignettes 5 (ITC_5_ = 0.634), 2 (ITC_2_ = 0.586), 4 (ITC_4_ = 0.504), as the other vignettes had a lower ITC with ITC_1_ = 0.307 and ITC_3_ = 0.110). The refined vignette set for SAAs can be found in the Appendix.

For LLMs, we identified nine vignettes with an ID of zero and seven vignettes with a negative ITC in step 1, all of which must be excluded, see Figure 1. However, only six vignettes have a positive ITC and for the remainder of the vignettes, an ITC could not be determined. Given the high number of exclusions and the inability to assess ITC for many vignettes, refining the vignette set for LLMs is unfeasible. Consequently, the full set must be retained for future evaluations.

### Comparing the Refined with the Original Vignette Set

The metrics obtained for each person using the refined vignette set showed a very high correlation with metrics obtained using the original vignette set, see Table 2. Similarly, the metrics obtained for each SAA using the refined vignette set showed very high correlations with metrics obtained using the original vignette set, see Table 3.

**Table 2.**
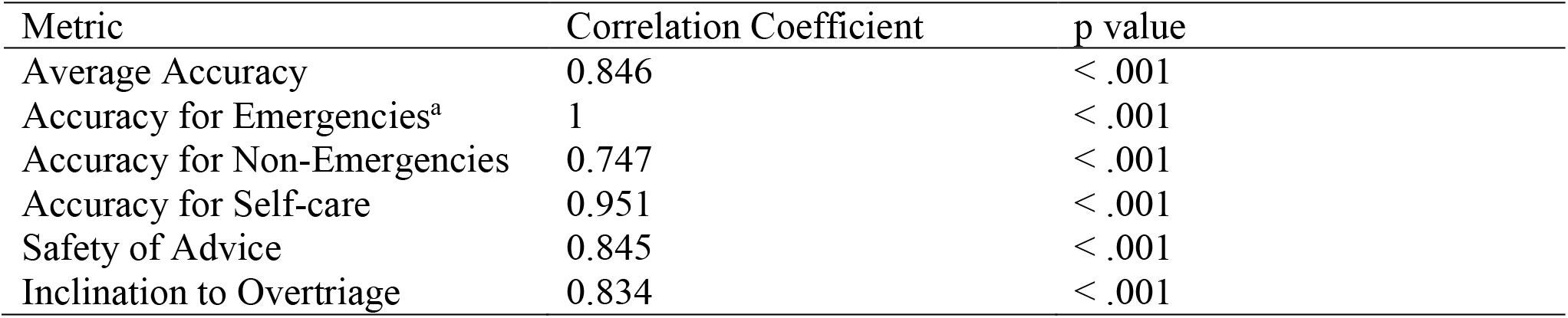
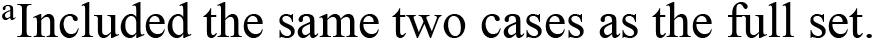
Correlations of metrics for laypeople for the refined versus the original vignette set.

| Metric | Correlation Coefficient | p value |
| --- | --- | --- |
| Average Accuracy | 0.846 | < .001 |
| Accuracy for Emergencies<sup>a</sup> | 1 | < .001 |
| Accuracy for Non-Emergencies | 0.747 | < .001 |
| Accuracy for Self-care | 0.951 | < .001 |
| Safety of Advice | 0.845 | < .001 |
| Inclination to Overtriage | 0.834 | < .001 |

<sup>a</sup>Included the same two cases as the full set.

**Table 3.** Correlations of metrics for SAAs for the refined versus the original vignette set.

| Metric | Correlation Coefficient | p value |
| --- | --- | --- |
| Average Accuracy | 0.995 | < .001 |
| Accuracy for Emergencies<sup>a</sup> | 0.577 | .04933 |
| Accuracy for Non-Emergencies | 0.989 | < .001 |
| Accuracy for Self-care | 0.986 | < .001 |
| Safety of Advice | 0.970 | < .001 |
| Inclination to Overtriage | 0.974 | < .001 |
| Capability Comparison Score | 0.997 | < .001 |

<sup>a</sup>Included only one out of two original cases. Correlation is unreliable, because estimates are either 1 or 0.

## Discussion

Overall, our analysis demonstrates that refining a vignette set as outlined in this study proves feasible. Results using the refined set show a very high correlation with performance based on the complete original set, indicating minimal loss of predictive power despite using fewer vignettes. This approach not only makes evaluations more cost-efficient by using fewer vignettes, but also ensures that only vignettes accurately predicting overall performance are included, thereby yielding more reliable performance estimates. This effort aligns with the call for standardized vignettes and their refinement (10–12,24). Answering Painter et al.’s call for guidance on which vignettes to include in triage evaluation studies (11), our two-step procedure offers a systematic, theory-driven way to refine an initial set of vignettes and select the most predictive vignettes out of a full set.

In our data, the ID was less relevant for refining vignettes for laypeople and SAAs, because at least one person or SAA managed to solve each case. The ID proved more meaningful for LLMs, however, which could rarely solve the self-care cases. This thwarted the refinement process for LLMs, because an ITC value was impossible to calculate for those vignettes that could not be solved by LLMs. In the current dataset, this problem might be due to the small number of LLMs (only five) included and the resulting low variance. With a higher number of LLMs, a refinement might be possible, but the number of different LLMs is currently limited.

The generated vignette sets vary between laypeople and SAAs. That is, a set validated for SAAs might not be suitable for assessing laypeople’s performance. So, researchers must refine the vignette set and collect data to validate it for each agent they wish to generalize to. Specifically, research should initially collect data using the full set and then refine and validate a subset to be used in follow-up studies with the same agent.

Our study comes with limitations that should be addressed in future studies. We focused on classical test theory to refine our vignette set, but item-response theory (IRT) could offer an alternative theoretical framework for the refinement. However, most models would require bigger sample sizes, which are often not available due to the limited number of SAAs (25). Until larger samples of SAAs become available, test theory is the best choice for refining vignettes for these agents. For laypeople, where larger sample sizes are feasible, exploring whether IRT yields different refined vignette sets could be a valuable next step. Additionally, assessing the validity (e.g., convergent and divergent validity) of case vignette sets presents a further research opportunity.

## Conclusions

Our two-step vignette refinement procedure offers a significant advancement for the evaluation of triage decision-making and digital health research at large. By systematically excluding vignettes that are unsuitable for measuring the constructs (e.g., triage accuracy) researchers wish to assess, they can avoid arbitrary selection of vignettes and ensure that only statistically validated vignettes are included in the test set. This approach can enhance the quality and reduce the costs of digital health research, lead to more reliable results, and enable more precise inferences in the long run. If more researchers apply the presented refinement method, methodological rigor and research quality will increase, which helps move the field forward and ultimately contributes to the development of more effective digital health tools and interventions.

## Data Availability

All data are available online at

https://zenodo.org/doi/10.5281/zenodo.12805048

